# Evaluation of risk stratification at presentation using the Alinity high-sensitivity cardiac troponin I assay

**DOI:** 10.64898/2026.08.29.26361405

**Authors:** Ziwen Li, Takeshi Fujisawa, Øyvind Skadberg, Paul Fineran, Alexander JF Thurston, Yong Yong Tew, Kristin M. Aakre, Nicholas L Mills, Ryan Wereski, the POC-ET Investigators

## Abstract

**Background:** High-sensitivity cardiac troponin (hs-cTn) assays enable safe early discharge of patients at very low risk for myocardial infarction. We previously developed a single-sample rule-out pathway using the ARCHITECT hs-cTnI assay to risk stratify patients with suspected acute coronary syndrome. In a secondary analysis of the POC-ET (Point of Care Evaluation of High-sensitivity Cardiac Troponin) study, we evaluated performance of risk stratification with the Alinity hs-cTnI assay.

**Methods:** Patients presenting with possible myocardial infarction in the POC-ET (NCT05665127) study were included. The primary outcome was type 1, 4b or 4c myocardial infarction or cardiac death at 30 days. Cardiac troponin I (cTnI) was measured in stored materials using the ARCHITECT and Alinity hs-cTnI assays. The sex-specific 99^th^ percentile upper reference limit (URL) are 34 ng/L in men and 16 ng/L in women for both assays. Agreement was assessed with Bland-and-Altman limit of agreement method, Passing Bablok regression, and Pearson’s correlation coefficient. Distributions of presentation measurements were compared with Kolmogorov-Smirnov test. Performance was evaluated in the overall population and prespecified subgroups. The negative predictive value (NPV) and sensitivity were determined and proportion of patients identified as low, intermediate, and high risk were calculated and modelled using ordinal logistic regression.

**Results:** In 986 patients (60 [51-70] years, 38% female), 78 (7.9%) had a primary outcome. Strong agreement was found in the raw cTnI measurements (99% samples within the Bland-Altman limit of agreement; correlation coefficient: 0.967 (95% CI 0.964-0.969, *P<0.001*); Passing Bablok regression: slope 1.12 [1.11-1.13], intercept-0.16 [-0.18 to-0.13]). At presentation, distributions of cTnI measurements by the two assays were similar (*P=0.810*). Both assays showed comparable diagnostic performance using a risk stratification threshold of <5 ng/L and the sex-specific diagnostic threshold, with the same NPV (Alinity 100 [99.7-100]% *versu*s ARCHITECT 100 [99.7-100]%) and sensitivity (Alinity 100 [97.3-100]% *versus* ARCHITECT 100 [97.3-100]%). Similar proportions of patients stratified as low-(Alinity 67% *versus* ARCHITECT 67%), intermediate-risk (23% *versus* 24%) and high-risk (10% *versus* 9%) at presentation with minor reclassification. Similar efficacy was observed across subgroups stratified by sex, age, history of myocardial infarction, renal function, and symptom duration.

**Conclusions:** The Alinity hs-cTnI and the ARCHITECT hs-cTnI assays can be used interchangeably in the assessment of suspected myocardial infarction with comparable safety and efficacy.

## Introduction

High-sensitivity cardiac troponin (hs-cTn) assays with enhanced precision at very low concentrations have enabled early detection of myocardial injury and improved efficiency in assessing patients presenting to the Emergency Department with suspected acute coronary syndrome.^1,2^ We previously developed a single-sample rule-out pathway for acute myocardial infarction using risk stratification and diagnostic thresholds derived from the Abbott ARCHITECT hs-cTnI assay.^3,4^ In a prospective, stepped-wedge, cluster randomized controlled trial HiSTORIC (High-Sensitivity Cardiac Troponin on Presentation to Rule Out Myocardial Infarction), we showed that the implementation of this pathway in clinical practice was safe and effective, reducing length of stay and hospital admission without compromising diagnostic performance.^5^ This study aimed to assess the agreement between the Abbott Alinity hs-cTnI assay and the ARCHITECT assay, and to evaluate the performance of risk stratification using the Alinity hs-cTnI assay in patients with suspected acute coronary syndrome.

## Methods

### Study design and population

This is a secondary analysis of the POC-ET (**P**oint **O**f **C**are **E**valuation of high-sensitivity cardiac **T**roponin) observational cohort study (clinicaltrials.gov registration number NCT05665127) of patients presenting with symptoms of suspected acute coronary syndrome presenting to the Emergency Department or Acute Medical Unit at three acute care hospitals in Scotland in whom the attending clinician requested high-sensitivity cardiac troponin. The inclusion criteria for POC-ET were: (1) presenting to hospital with symptoms of suspected acute coronary syndrome; (2) age 18 years or over. Patients were excluded if they: (1) ST-segment elevation on the electrocardiogram; (2) presented with an out-of-hospital cardiac arrest; (3) were unable or unwilling to give informed consent; (4) were unable or unwilling to comply with study protocol; (5) previously enrolled in the study. Patients were eligible for inclusion in this secondary analysis if plasma or serum samples were available and of suitable quality for analysis. Patients were identified as low risk if they presented after 2 hours from symptom onset and had cardiac troponin concentrations below 5 ng/L at presentation, and as high risk if cardiac troponin concentrations were above the sex-specific 99^th^ percentile upper reference limit. Patients who had cardiac troponin concentrations between 5 ng/L and the sex-specific 99^th^ percentile at presentation or presented within 2 hours of symptom onset were identified as intermediate risk. Individual patient consent was obtained from all participants, and all data were deidentified and linked prior to being made available to the research team. The study was approved by the Health and Social Care Research Ethics Committee B (Reference: 21-NI-0200) and was conducted according to the principles of the Declaration of Helsinki.

### Cardiac troponin testing

Plasma or serum samples collected at 0 hr, 1 hr, 2 hr, and 6-36 hours after presentation were stored at the BHF Cardiac Biomarker Laboratory at below-70°C prior to measurement of hs-cTnI.

Troponin I concentrations were measured in batches after completing the recruitment at the all participating sites using the ARCHITECT high-sensitivity cardiac troponin I assay (Abbott Diagnostics) in the lithium heparin plasma samples on the ARCHITECT ci4100 platform at the BHF Cardiac Biomarker Laboratory, the University of Edinburgh. The ARCHITECT assay has an inter-assay coefficient of variation of less than 10% at 4.7 ng/L. The uniform 99^th^ percentile upper reference limit is 26 ng/L, and the sex-specific 99^th^ percentile is 34 ng/L in men and 16 ng/L in women.^6^

Serum samples were transferred to the Laboratory of Medical Biochemistry, Stavanger University Hospital, and stored at below-70°C prior to measurement of hs-cTnI on the Abbott Alinity platform. This assay has an inter-assay coefficient of variation of less than 10% at 4.7 ng/L. The uniform 99^th^ percentile upper reference limit is 26 ng/L, and the sex-specific 99^th^ percentile is 34 ng/L in men and 16 ng/L in women.^6^

All samples were analysed by trained laboratory technicians in accordance with the manufacturer’s package insert.

### Data collection and adjudication

Baseline and outcome data were recorded in an electronic study database (Research Electronic Data Capture, Vanderbilt University).^7^ The final diagnosis was adjudicated by two independent clinicians in all patients with evidence of myocardial injury (cardiac troponin concentration greater than sex-specific 99^th^ centile upper reference limit) within 24 hours of index presentation using the ARCHITECT hs-cTnI assay. Where there was disagreement a third clinician arbitrated. Adjudication was based on the Fourth Universal Definition of Myocardial Infarction^1^ following review of all clinical and laboratory information available to the usual care team. The adjudicators were blinded to the Alinity hs-cTnI assay results Follow-up data were collected by site investigators and uploaded to Secure Data Environment DataStore at the University of Edinburgh for analysis.

## Statistical analysis

We assessed the agreement in raw hs-cTnI measurements using the Bland-and-Altman limits of agreement method and regression models including Passing Bablok regression (non-parametric method), Deming regression (allowing uncertainty on both x and y axes), and simple linear regression (allowing uncertainty on the y axis), and calculated the Pearson’s correlation coefficient. We compared the distribution of rounded hs-cTnI measurements at presentation using a two-sample Kolmogorov-Smirnov test, and evaluated the difference in risk stratification group-allocation at presentation using an ordinal logistic regression. Alluvial plots were used to illustrate reclassification. We determined the negative predictive value (NPV), positive predictive value (PPV), sensitivity, and specificity for the primary outcome of myocardial infarction (type 1, type 4b or type 4c myocardial infarction) or cardiac death during the index presentation or following discharge at 30 days. The 95% confidence intervals (CI) were estimated using a Bayesian binomial inference with Jeffreys prior to ensure optimal coverage for NPV and sensitivity that were expected to approach 100%. The diagnostic performance was evaluated in the total study population and in prespecified subgroups stratified by age, sex, time since symptom onset, history of previous myocardial infarction, and by renal function (eGFR). All analyses were conducted in R version 4.5.3 (R Foundation for Statistical Computing).

## Results

### Cardiac troponin measurements using the Alinity and ARCHITECT hs-cTnI assays

We assessed 2,848 blood samples from 988 patients with suspected acute coronary syndrome from the POC-ET observational study cohort using the Alinity and the ARCHITECT hs-cTnI assays. After excluding those with insufficient sample volume the analysis was performed on 2,834 samples from 986 patients (***Table 1***). Raw measurements obtained using the ARCHITECT assay had a median of 3.3 ng/L (interquartile range (IQR) 7.5 ng/L). Measurements obtained using the Alinity assay had a median of 3.4 ng/L (IQR 7.5 ng/L). We assessed agreement in paired measurements obtained using the two assays from the same sample, using the Bland and Altman limits of agreement method^8^ and three regression models. Each data point in the Bland-Altman plot (***Figure 1***) represented a sample, and 99% (2,808/2,834) samples were within the Bland and Altman limits of agreement (LoA) defined as Measurements of 26 samples from 17 individual patients were outside of the LoA, among whom 14 were males.

**Figure 1.**
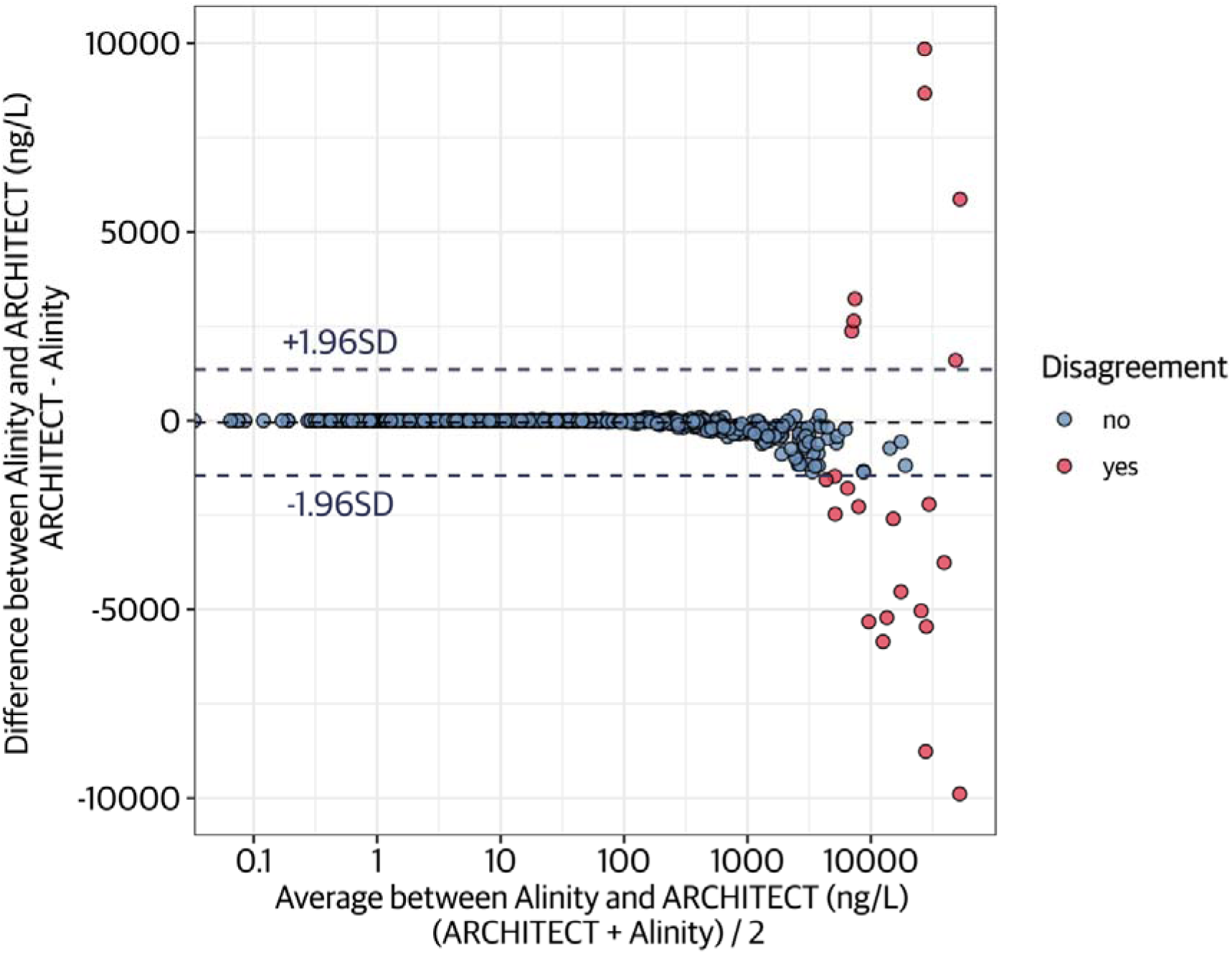
Bland-Alman plot to evaluate the agreement between hs-cTnI measurements by Alinity and ARCHITECT. The data points were coloured by agreement.

**Table 1.** Baseline characteristics.

| Baseline characteristics | n = 986 |
| --- | --- |
| Age | 60 (51, 70) |
| Male | 609 (62) |
| Primary outcome | 78 (7.9) |
| Presentation complaint |  |
| Chest pain | 930 (94) |
| Dyspnoea | 17 (1.7) |
| Palpitation | 12 (1.2) |
| Other | 27 (2.7) |
| Myocardial ischaemia on ECG | 201 (20) |
| Family history | 352 (36) |
| Previous medical conditions |  |
| Hypertension | 364 (37) |
| Diabetes mellitus | 150 (15) |
| Angina | 186 (19) |
| Myocardial infarction | 210 (21) |
| Heart failure | 42 (4.3) |
| Stroke | 57 (5.8) |
| Peripheral vascular disease | 20 (2.0) |
| Previous revascularisation |  |
| PCI | 185 (19) |
| CABG | 28 (2.8) |
| Medication at presentation |  |
| Statin | 431 (44) |
| Aspirin | 258 (26) |
| ACEI or ARB | 317 (32) |
| Beta blocker | 270 (27) |
| Oral anti-coagulant | 95 (9.6) |
| Clinical chemistry |  |
| eGFR, mL/min | 93 (79, 102) |
| Presentation hs-cTnI (Alinity), ng/L | 3 (2, 8) |
| Presentation hs-cTnI (ARCHITECT), ng/L | 3 (2, 8) |

Values are n (%) or median (Q1-Q3). ECG, electrocardiogram; PCI, percutaneous coronary intervention; CABG, coronary artery bypass grafting; ACEI, angiotensin-converting enzyme inhibitor; ARB, angiotensin receptor blocker; eGFR, estimated glomerular filtration rate; hs-cTnI, high-sensitivity cardiac troponin I.

Pearson’s correlation coefficient was calculated and three regression models (***Figure 2***), Passing Bablok regression, Deming regression, and simple linear regression, were used to evaluate the correlation in the measurements by the two assays. The Pearson’s correlation coefficient was 0.967 (95% CI 0.964-0.969, *P<0.001*). The distributions of hs-cTnI measurements by the two assays were right-skewed and the non-parametric Passing Bablok regression was the most suitable model, which gave a slope of 1.12 (95% CI 1.11 to 1.13), close to 1, and an intercept of-0.16 (95% CI-0.18 to-0.13), close to 0. For measurements within the range of 0 to 1000 ng/L, the Pearson’s correlation coefficient was 0.980 (95% CI 0.978-0.981, *P<0.001*), and the slope and intercept of the Passing Bablok regression model were 1.10 (95% CI 1.09 to 1.11) and-0.12 (95% CI-0.14 to-0.09). The other two regression models, assuming normal distribution of underlying data, were included for reference. The majority of samples had small absolute difference between paired measurements (***Table 2***), and strong agreement was observed at the lower end of the measurable range, including the risk stratification threshold of <5 ng/L and the sex-specific 99^th^ percentile upper reference limit (16 ng/L in women and 34 ng/L in men), used to inform risk stratification in the Emergency Department (***Figure 3***).

**Figure 2.**
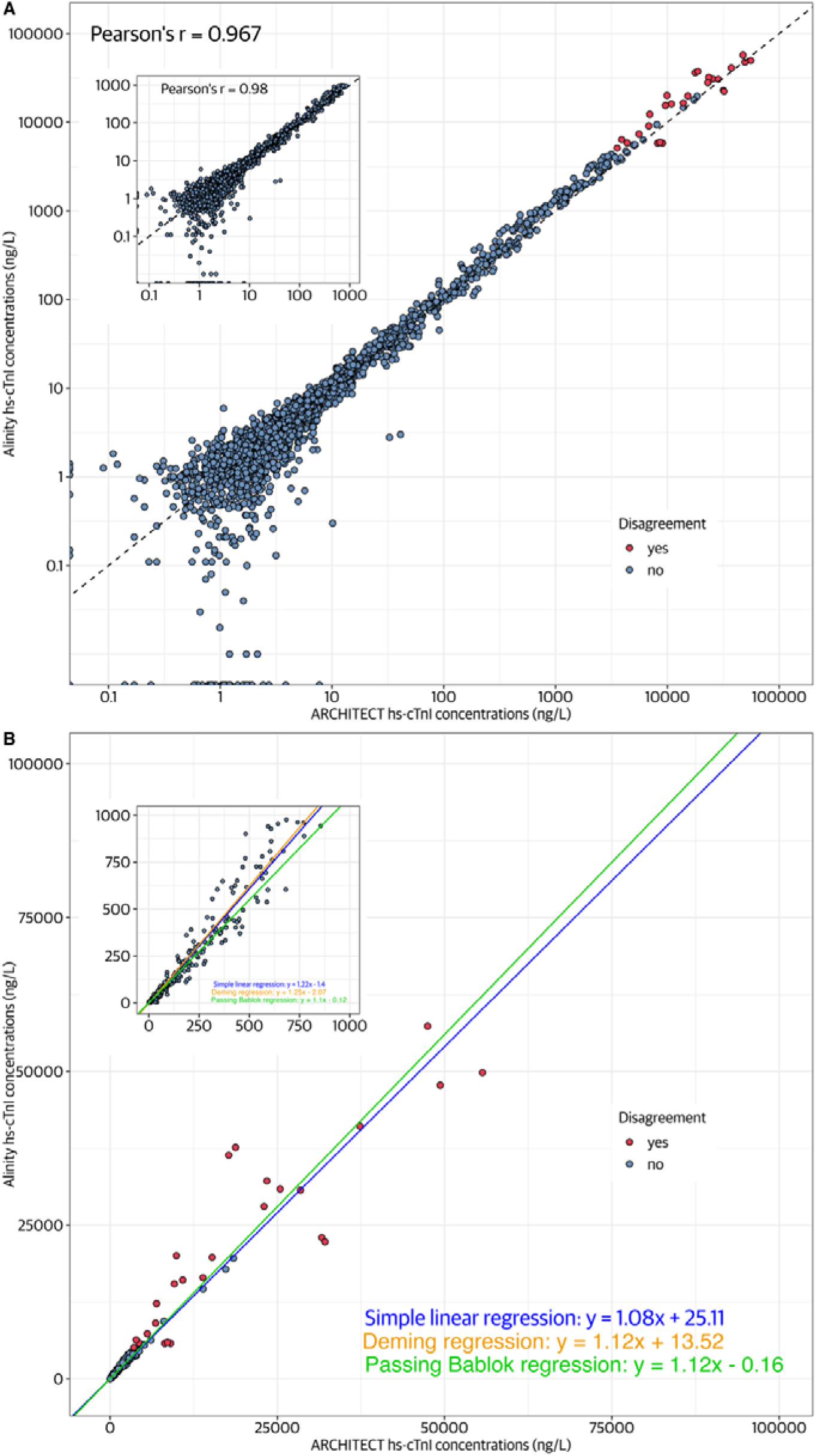
Scatter plots to visualise the agreement between hs-cTnI measurements by Alinity and ARCHITECT. The data points were coloured by agreement based on the Bland-Altman limits of agreement analysis. (A) Scatter plot with both axes on log10 scales. The equality line y = x was plotted (dashed line) to provide reference for perfect agreement. (B) Scatter plot with both axes on normal scales. Three regression lines were plotted for the three simple regression models, simple linear regression (blue), Deming regression (orange), and Passing Bablok regression (green). The estimates and 95% CI for the slopes and intercepts were presented in Table 1. The plots within plots A and B were for values within the range of 0-1000 ng/L for both Alinity and ARCHITECT.

**Figure 3.**
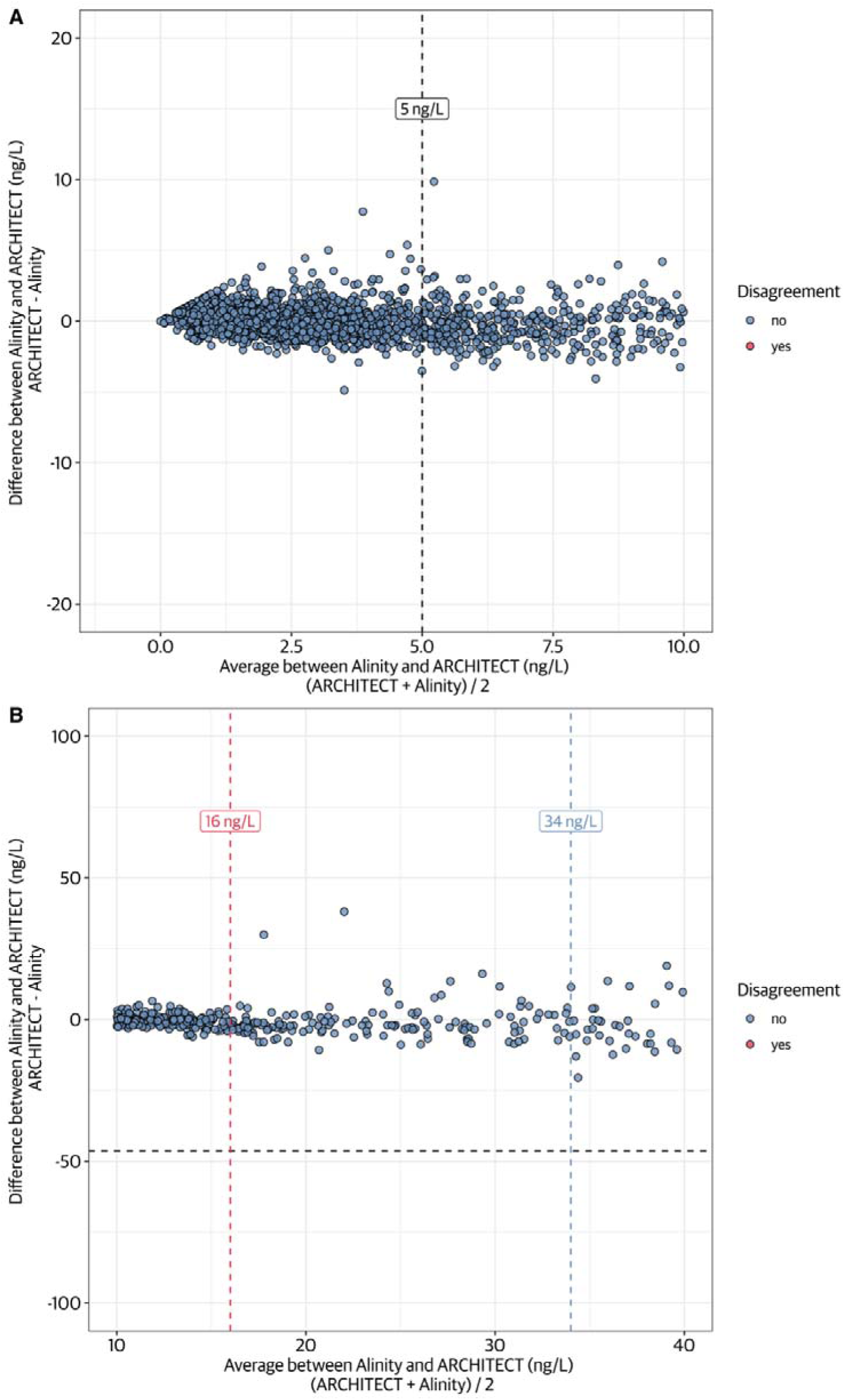
Scatter plots to visualise the agreement between hs-cTnI measurements by Alinity and ARCHITECT at lower end of measurable range. The data points were coloured by agreement based on the Bland-Altman limits of agreement analysis. Risk stratification threshold of <5 ng/L and sex-specific 99^th^ percentile upper reference limit (16 ng/L for women and 34 ng/L for men) were indicated using dashed vertical lines. The mean difference was indicated with a dashed horizontal line.

**Table 2.** Counts and percentage of samples with small absolute difference using the STAT High Sensitive Troponin-I assays on the Alinity and the ARCHITECT (ng/L).

| **Difference** | **<1 ng/L** | **<2 ng/L** | **<3 ng/L** | **<4 ng/L** | **<5 ng/L** | **<6 ng/L** | **<7 ng/L** |
| --- | --- | --- | --- | --- | --- | --- | --- |
| Counts | 1,730 | 2,221 | 2,382 | 2,447 | 2,481 | 2,501 | 2,512 |
| Percentage | 61% | 78% | 84% | 86% | 88% | 88% | 89% |

Distributions of hs-cTnI concentrations at presentation measured by the Alinity and the ARCHITECT assays were visualised using density plots (***Figure 4***), and compared using a two-sample Kolmogorov-Smirnov test (*P=0.810*), with similar distribution.

**Figure 4.**
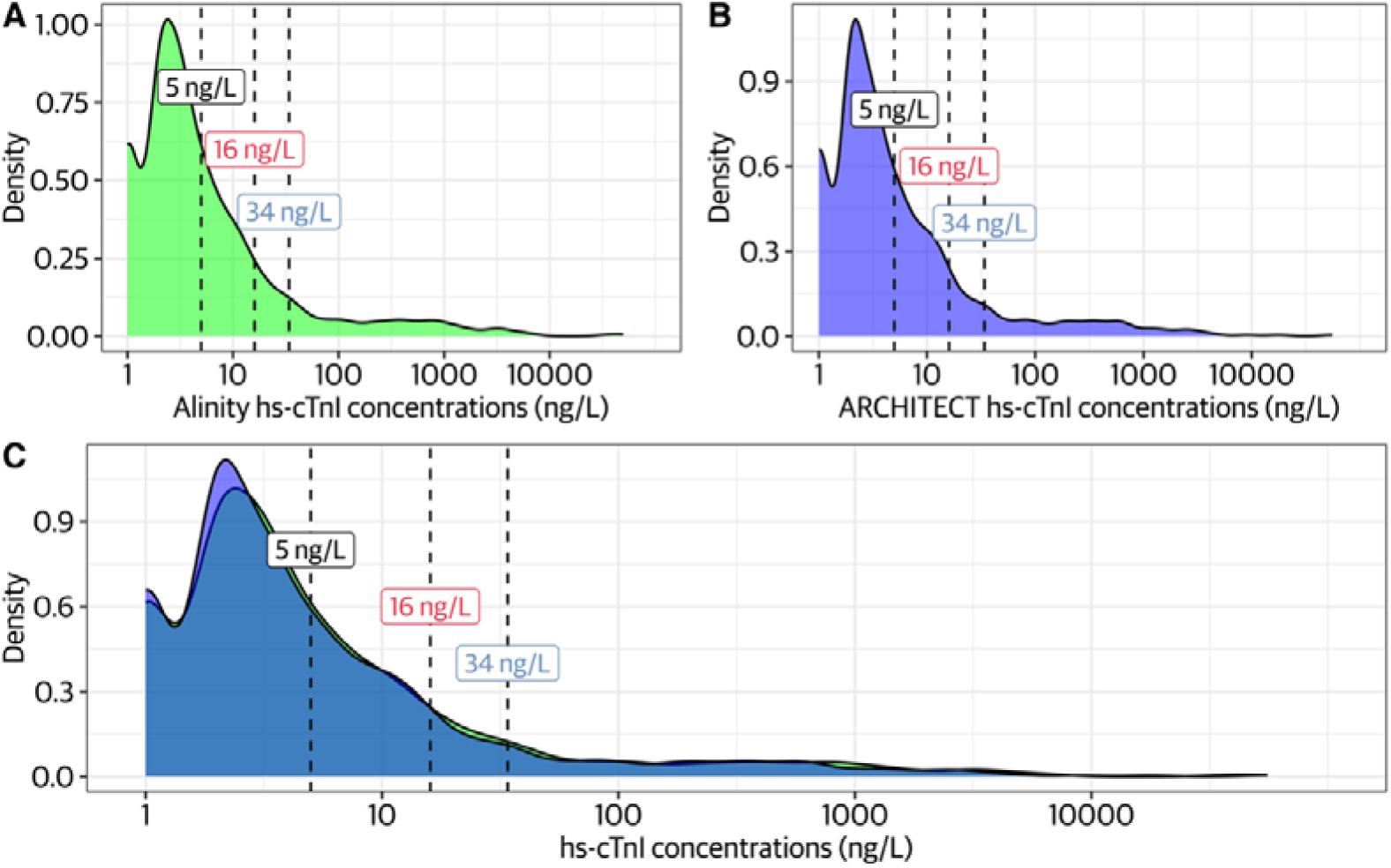
Distribution of hs-cTnI concentrations at presentation measured by Alinity (A) and ARCHITECT (B). (C) is overlay of A and. **B.** Optimal risk stratification threshold (5 ng/L) and sex-specific 99^th^ URLs (16 ng/L for women and 34 ng/L for men) were indicated using dashed lines.

### Efficacy of risk stratification at presentation

The efficacy of risk stratification using hs-cTnI concentrations at presentation was assessed using the established risk stratification threshold of <5 ng/L and the sex-specific 99^th^ percentile upper reference limit in 876 patients with presentation measurements using both assays. Ischaemic findings on the electrocardiogram (ECG) were observed in 16.2% (142/876) patients at presentation. Analyses of efficacy and reclassification were performed in the 734 patients without ischaemic ECG findings. Alluvial plots were used to visualise reclassification using the two assays at presentation (***Figure 5***).

**Figure 5.**
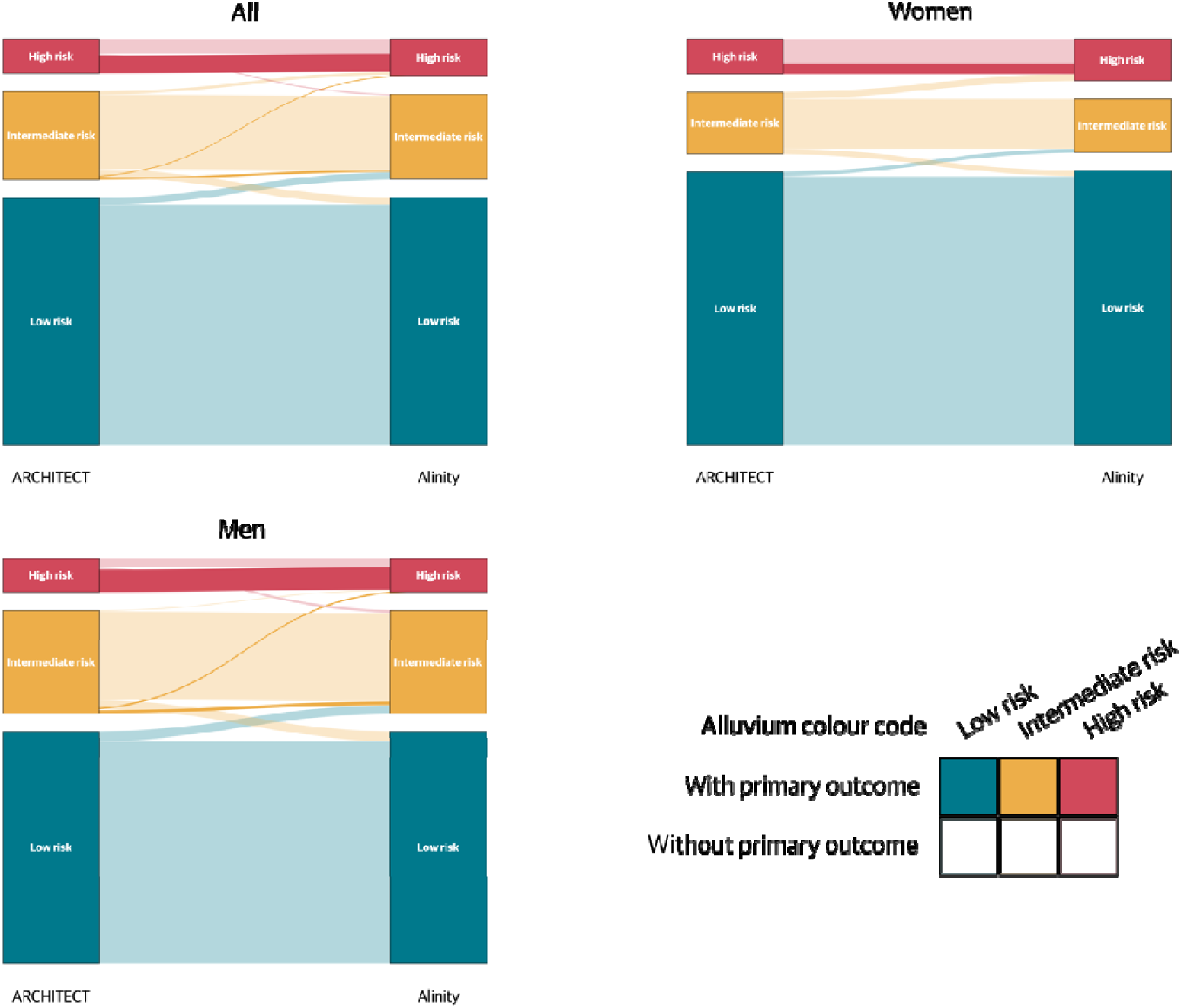
Reclassification of patients using the Alinity hs-cTnI assay at presentation in all patients and by sex. Patients identified as low, intermediate, and high risk were indicated using green, yellow, and red colours, and darker hues indicated individuals with the primary safety outcome.

Using the ARCHITECT hs-cTnI assay, 66.9% (491/734) patients were identified as low risk, 23.8% (175/734) as intermediate risk for retest, and 9.3% (68/734) as high risk. In comparison, using the Alinity assay, 67.0% (492/734) patients were identified low risk, 23.0% (169/734) as intermediate risk, and 9.9% (73/734) as high risk. Of the patients classified as low-risk using the ARCHITECT hs-cTnI assay, 2.9% (14/491; 3 females and 11 males) were re-classified as intermediate risk using the Alinity assay. In contrast, of the patients classified as intermediate risk on the ARCHITECT, 8.6% (15/175; 4 females and 11 males) and 4.6% (8/175; 5 females and 3 males) were re-classified as low and high risk, respectively. A small number of patients classified as high risk on the ARCHITECT hs-cTnI assay were re-classified as intermediate risk (4.4% (3/68; 3 males)). No high-risk patient identified using the ARCHITECT hs-cTnI assay were reclassified as low risk using the Alinity assay. No patients reclassified by the Alinity assay from a higher to a lower risk group had a primary outcome event.

Ordinal logistic regression models were fitted to compare risk stratification group-allocation using the two assays. This showed that the probabilities of low-, intermediate-, and high-risk patients identified using the ARCHITECT assay, who would remain in the same risk group when switching to the Alinity assay, were 96%, 86%, and 96%. When stratified by sex, the corresponding probabilities were 98%, 81%, and 100% in women, and 94%, 88%, and 93%, in men.

### Diagnostic performance at presentation

The performance of risk stratification at presentation was assessed using the Alinity hs-cTnI assay compared to the ARCHITECT hs-cTnI assay, for the primary outcome of myocardial infarction (type 1, type 4b or type 4c myocardial infarction) or cardiac death from index to 30 days. The impact of each assay on the negative predictive value (NPV), positive predictive value (PPV), sensitivity, and specificity, were evaluated using the risk stratification threshold of <5 ng/L alone or in conjunction with ischaemic ECG findings at presentation (***Table 3***).

**Table 3.** Safety metrics for using the risk stratification threshold of < 5 ng/L in all patients and those without ischaemic ECG findings at presentation.

|  | All patients |  | Non-ischaemic ECG patients |  |
| --- | --- | --- | --- | --- |
|  | Alinity | ARCHITECT | Alinity | ARCHITECT |
| **Counts** |  |  |  |  |
| Total | 876 | 876 | 734 | 734 |
| True Positive | 69 | 69 | 42 | 42 |
| True Negative | 562 | 564 | 492 | 491 |
| False Positive | 245 | 243 | 200 | 201 |
| False Negative | 0 | 0 | 0 | 0 |
| **Metrics (95% CI)** |  |  |  |  |
| NPV, % | 100 (99.7,100) | 100 (99.7, 100) | 100 (99.6, 100) | 100 (99.6, 100) |
| Sensitivity, % | 100 (97.3,100) | 100 (97.3, 100) | 100 (95.6, 100) | 100 (95.6, 100) |
| PPV, % | 22.0 (17.7, 26.8) | 22.1 (17.8, 27.0) | 17.4 (13.0, 22.5) | 17.3 (12.9, 22.4) |
| Specificity, % | 69.6 (66.4, 72.7) | 69.9 (66.7, 73.0) | 71.1(67.6, 74.4) | 71.0 (67.5, 74.2) |

When excluding patients with ischaemic ECG findings at presentation, a rule-out threshold of <5 ng/L had an NPV of 100 (95% CI, 99.6-100%) and a sensitivity of 100 (95% CI, 95.6-100%) using both assays. When all patients were assessed, the risk stratification threshold of <5 ng/L gave an NPV of 100 (95% CI, 99.7-100%) and a sensitivity of 100 (95% CI, 97.3-100%) using both the Alinity and the ARCHITECT hs-cTnI assays.

### Subgroup analysis

The proportions identified as low, intermediate, and high risk using the Alinity and the ARCHITECT hs-cTnI assays were similar in pre-specified subgroups stratified by sex, age, previous myocardial infarction, renal function, and time from symptom onset (***Table 4***).

**Table 4.** Efficacy of risk stratification using the ARCHITECT and the Alinity in pre-specified groups.

|  |  | ARCHITECT, count (%) |  |  | Alinity, count (%) |  |  |
| --- | --- | --- | --- | --- | --- | --- | --- |
|  |  | Low risk | Intermediate risk | High risk | Low risk | Intermediate risk | High risk |
| **Sex** |  |  |  |  |  |  |  |
|  | men (n = 458) | 287 (63) | 129 (28) | 42 (9) | 287 (63) | 129 (28) | 42 (9) |
|  | women (n = 276) | 204 (74) | 46 (17) | 26 (9) | 205 (74) | 40 (14) | 31 (11) |
| **Age** |  |  |  |  |  |  |  |
|  | >65 years (n = 240) | 116 (48) | 87 (36) | 37 (15) | 114 (48) | 90 (38) | 36 (15) |
|  | ≤65 years (n = 494) | 375 (76) | 88 (18) | 31 (6) | 378 (77) | 79 (16) | 37 (7) |
| **Previous MI** |  |  |  |  |  |  |  |
|  | yes (n = 151) | 77 (51) | 53 (35) | 21 (14) | 73 (48) | 56 (37) | 22 (15) |
|  | no (n = 583) | 414 (71) | 122 (21) | 47 (8) | 419 (72) | 113 (19) | 51 (9) |
| **eGFR** |  |  |  |  |  |  |  |
|  | <60 mL/min (n = 71) | 23 (32) | 35 (49) | 13 (18) | 24 (34) | 34 (48) | 13 (18) |
|  | ≥60 mL/min (n = 663) | 468 (71) | 140 (21) | 55 (8) | 468 (71) | 135 (20) | 60 (9) |
| **Early presenter\*** |  |  |  |  |  |  |  |
|  | yes (n = 124) | 91 (73) | 23 (19) | 10 (8) | 89 (72) | 22 (18) | 13 (10) |
|  | no (n = 610) | 400 (66) | 152 (25) | 58 (10) | 403 (66) | 147 (24) | 60 (10) |

\* Early presenters are defined as those presented within 2 hours since symptom onset.

## Discussion

In this secondary analysis of the multi-centre observational cohort study POC-ET, we assessed the agreement in cardiac troponin measurements by the Alinity hs-cTnI assay and the ARCHITECT hs-cTnI assay in patients presenting to the Emergency Department with suspected acute coronary syndrome. High degree of agreement was found in measurements obtained using the two assays, especially at the lower end of the measurable range used to guide patient care and inform risk stratification. Similar proportions of patients were stratified as low, intermediate, and high risk at presentation, with minor re-classification. There was similarly good safety performance (NPV 99.5% and sensitivity 99%) between both assays. No patients reclassified from a higher to a lower risk group using the Alinity assay had a primary outcome event. Efficacy was comparable across subgroups stratified by sex, age, previous myocardial infarction, renal function, and time since symptom onset. These findings suggest that the Alinity assay and the ARCHITECT assay can be used interchangeably in the assessment of suspected myocardial infarction with excellent efficacy and safety.

Our study has several strengths. Paired measurements of cardiac troponin were obtained from a contemporary cohort of patients with suspected acute coronary syndrome using the Alinity and the ARCHITECT hs-cTnI assays. Multiple methods were used to evaluate agreement between the two assays. Diagnoses were adjudicated using measurements from the gold standard ARCHITECT hs-cTnI assay. Established rule-out threshold and guideline recommended sex-specific diagnostic thresholds were employed to assess the performance of risk stratification at presentation using both assays.

Our study also has important limitations. The blood samples submitted for testing were prepared differently: serum samples were used for Alinity testing and lithium heparin samples were used for ARCHITECT, and it is unclear whether using identical sample preparation methods would result in better or worse agreement in raw measurements or clinical performance. Both sample types can be used in practice in line with manufacturers recommendations. In addition, the number of events was modest and additional validation in cohorts with higher event rates would be useful to ensure generalisability. Finally, this was a retrospective study using stored blood samples, and did not assess the impact of assay implementation in practice.

## Conclusion

The Alinity hs-cTnI and the ARCHITECT hs-cTnI assays can be used interchangeably in the assessment of suspected myocardial infarction with comparable safety and efficacy.

## Author Contributors

RW, TF, and NLM conceived the study and its design. ZL, TF, ØS, and RW performed the analysis. ZL, TF, RW and NLM interpreted the data. ZL, TF, RW, and NLM drafted the manuscript. ZL, TF, ØS, PF, AJFT, YYT, KMA, RW, and NLM revised the manuscript critically for important intellectual content. All authors provided their final approval of the version to be published. All authors are accountable for the work.

## Authors’ Disclosures or Potential Conflicts of Interest

AJFT reports a research grant and speaker honorarium received from Roche Diagnostics outside the submitted work. KMA has served on advisory boards for Roche Diagnostics, Abbott Diagnostics, Siemens Healthineers, and SpinChip; received consultancy honoraria from CardiNor; lecture honoraria from Siemens Healthineers, Roche Diagnostics, Mindray, Wondfo, and Snibe Diagnostics; and research grants from Siemens Healthineers and Roche Diagnostics. KMA is an Associate Editor of Clinical Biochemistry and Chair of the IFCC Committee on Clinical Application of Cardiac Biomarkers. NLM has received honoraria or consultancy from Abbott Diagnostics, Roche Diagnostics, and Siemens Healthineers within the last 36 months. RW has received insitional research grants from Novo Nordisk and Abbott Diagnostics. All other authors have reported that they have no relationships relevant to the contents of this paper to disclose.

## Funding

This study was supported by an Investigator Initiated Study award from Abbott Laboratories to the University of Edinburgh. ZL is supported by a British Heart Foundation Project Grant (PG/25/12298). AJFT is supported by a British Heart Foundation Clinical Research Training Fellowship (FS/CRTF/25/24635). YYT is supported by the British Heart Foundation Scholarship (CH/F/21/90010). NLM is supported by a Chair Award, Programme Grant, and Research Excellence Award (CH/F/21/90010, RG/F/25/110169, RE/24/130012) from the British Heart Foundation.

## Data Availability

All data produced in the present study are available upon reasonable request to the authors.

## Acknowledgements

We gratefully acknowledge the BHF Cardiovascular Biomarker Laboratory, the University of Edinburgh, Department of Clinical Biochemistry (Glasgow Royal Infirmary, NHS Greater Glasgow and Clyde), and the Stavanger University Hospital Laboratory of Clinical Biochemistry for their expertise and assistance in this work.

## POC-ET investigators

Atul Anand, Lynda Attwood, Jennifer Blades, Ross Campbell, Andrew R Chapman, Claire Cheyne, Jamie G Cooper, Amanda Coutts, Amy V Ferry, Paul Fineran, Takeshi Fujisawa, Alasdair J Gray, Deepak Harry, James Henderson, Elizabeth Highton-Williamson, Ziwen Li, Stephen Lynch, Fiona McCurrach, Michael McDermott, Shirley McDonald, Nicholas L Mills, Mizu Mizerska, David E Newby, Jennifer Noble, Rachel O’Brien, Judi O’Shaughnessy, Onyinyechi Oziogu, Salan Rowlands, Andrew Sorbie, Caelan Taggart, Yong Yong Tew, Alexander JF Thurston, Christopher Tuck, Daniel P Vicencio, Jess Walters, Ryan Wereski.

## Notes

### Clinical Trial

NCT05665127

### Author Declarations

The study was approved by the Health and Social Care Research Ethics Committee B (Reference:21-NI-0200).

